# RNA sequencing identifies human placental *IL3RA* as a potential predictor of adverse cardiovascular outcomes in patients with severe preeclampsia

**DOI:** 10.1101/2023.06.16.23291528

**Authors:** Omonigho Aisagbonhi, Tony Bui, Hailee St. Louis, Donald Pizzo, Morgan Meads, Megan Mulholland, Robert Morey, Celestine Magallanes, Leah Lamale-Smith, Louise C. Laurent, Marni B. Jacobs, Kathleen M. Fisch, Mariko Horii

## Abstract

**Background:** Mortality from preeclampsia (PE) and PE-associated morbidities are 3-to 5-fold higher in persons of African ancestry than in those of Asian and European ancestries. The placenta is central to the etiology of PE. However, how and to what extent the placenta contributes to worse PE outcomes in persons of African ancestry is yet to be fully elucidated.

**Objective:** We aimed to identify molecular pathways that are unique or enriched in placentas of parturient persons of African ancestry with PE with severe features (sPE) compared to those of Asian and European ancestry with sPE.

**Study design:** Bulk RNA sequencing was performed on 50 placentas from parturient persons with sPE of African (n=9), Asian (n=18) and European (n=23) ancestries and 73 normotensive controls of African (n=9), Asian (n=15) and European (n=49) ancestries.

**Results:** Metabolism, hormone regulation and hypoxia/angiogenesis genes, previously described to be upregulated in PE, including: *LEP*, *PAPPA2*, *INHA*, *FSTL3*, *FLT1, PHYHIP* and *ENG*, were upregulated in sPE across ancestries, with high expression of *FSTL3* being additionally associated with intrauterine growth restriction (p = .0047). Notably, the upregulation of, *FLT1*, *LEP* and *PHYHIP* was significantly higher in sPE placentas from parturient persons of African versus Asian ancestry (p = .0.35, .020 and .012 respectively). Genes associated with allograft rejection and adaptive immune response were upregulated in placentas from parturients of African ancestry but not in those of Asian and European ancestries. Among the allograft rejection/adaptive immune response genes, *IL3RA* was of particular interest because the patient with the highest placental *IL3RA* level, a woman of African ancestry with *IL3RA* levels 4.5-fold above the average for African ancestry parturients with sPE, developed postpartum cardiomyopathy, and was the only patient out of 123, that developed this condition. Interestingly, the sPE patients with the highest *IL3RA* levels among parturients of Asian and European ancestries developed unexplained tachycardia peripartum, necessitating echocardiography in the European ancestry patient. The association between elevated placental *IL3RA* levels and unexplained tachycardia or peripartum cardiomyopathy was found to be significant in the 50 sPE patients (p = .0005).

**Conclusions:** Placentas from parturients of African ancestry express higher levels of metabolism (*LEP*) and hypoxia/angiogenesis (*FLT1*) genes, as well as allograft rejection/adaptive immune response genes, including *IL3RA*. High placental expression of *IL3RA* may predict worse maternal cardiovascular outcomes, including peripartum cardiomyopathy. Studies evaluating placental *IL3RA* levels in peripartum cardiomyopathy cohorts are therefore warranted, as are broader studies evaluating placental factors in maternal cardiovascular outcomes postpartum.

## Introduction

Preeclampsia (PE) is a hypertensive disorder of pregnancy (HDP) that the WHO lists as one of the 5 major causes of maternal mortality worldwide^1^. Pregnant and parturient persons of African ancestry have 1.5 to 2.5-times higher rates of PE^2–5^ and are 3 to 5-times more likely to die of PE than pregnant and parturient persons of Asian or European ancestry^6–7^. PE-associated morbidities, such as, stroke, pulmonary edema and heart failure, are also significantly higher in pregnant and parturient persons of African ancestry^6–7^.

Underlying comorbidities and lower socioeconomic status (SES) do not entirely explain the higher PE rates and worse prognosis in persons of African ancestry. A population-based study of 718,604 Black and White pregnant and parturient persons from a California cohort of singleton births found that, although White persons of higher SES had a lower risk of PE than White persons of lower SES, higher SES did not attenuate PE risk in Black persons^8^. A review of the records of 1,355 pregnant persons with HDP found similar rates of underlying chronic hypertension in Black and White pregnant persons. However, among pregnant persons with HDP without underlying chronic hypertension, Black persons were significantly more likely to be diagnosed with PE than White persons^9^. In their retrospective cohort study of California births in 2007, Snowden et al^10^ found that though obesity was a risk factor for PE and low birth weight in White pregnant persons, obesity did not confer significantly increased PE risk in Black persons. However, Black pregnant persons of normal weight had a higher risk of PE than White persons of normal weight.

The placenta plays a central role in the pathology of PE, as PE only occurs in the context of pregnancy, and delivery of the placenta is often curative^11–13^. However, how the placenta contributes to worse PE outcomes in pregnant and parturient persons of African ancestry, and which specific placental injury pathways may be dysregulated in this population (and therefore potential targets for therapy), are not well studied.

Molecular and clinical data point to multiple pathways leading to PE. Using unsupervised clustering of aggregate microarray datasets (first in 173 patient samples, including 77 with PE and 96 controls, and later in a larger dataset of 330 samples, including 157 PE and 173 non-PE samples), Leavey et al^14–15^ identified 3 molecular clusters/subclasses of PE which they termed maternal, canonical and immunologic. In the maternal PE group, infant birthweight and placental weight were appropriate for gestational age, and placental histology and gene expression patterns were not significantly different from normotensive controls. In the canonical PE group, birthweight and placental weight were small for gestational age, placental histology showed evidence of maternal vascular malperfusion, and genes associated with hormone regulation and angiogenesis were overexpressed, with *FSTL3* identified as the top differentially expressed gene. In the immunologic PE group, birthweight and placental weight were small for gestational age, placental histology showed massive/increased perivillous fibrin deposition, and genes associated with immune and inflammatory responses were overexpressed.

Recently, our group performed RNA sequencing on placentas from PE patients with evidence of maternal vascular malperfusion and found trophoblast defects and activation of pathways associated with hypoxia, inflammation and reduced cell proliferation in placentas with maternal vascular malperfusion^16^, thus demonstrating that placental tissue RNA sequencing reflects pathways associated with histologic evidence of placental injury in PE.

Therefore, we sought to elucidate the cellular and molecular underpinnings of the worse PE outcomes in pregnant and parturient persons of African ancestry by performing bulk RNA sequencing on placentas from persons affected by PE with severe features (sPE) and normotensive controls of African ancestry compared to persons of Asian and European ancestries. We hypothesized that molecular PE subclasses differ in placentas from parturient persons of African ancestry versus those of Asian and European ancestries.

## Materials and Methods

### Study cohort selection

The study was approved by University of California, San Diego (UCSD) institutional review board, approval #181917. The study specimens were obtained from 2655 singleton parturitions at UCSD-affiliated hospitals between January 2010 and November 2020. The patients whose samples were collected gave written informed consent for accessing their electronic medical records and collection of placental tissue. Demographic, clinical and placenta histology data were collected in a REDCap-based obstetric registry. For each pregnancy, the clinical diagnosis of preeclampsia with severe features (sPE) was adjudicated by 2 Board-Certified obstetricians. Of the 2655 parturitions, 1003 for which the maternal ethnicity was recorded as Hispanic-other were excluded, because Hispanic is a language group that can include people of diverse ancestries, ranging from African (e.g. Afro-Cubans) to North and South Americans (e.g. Mexicans and Brazilians), Asians (e.g. Spanish Filipinos) and Europeans (e.g. Spaniards). Categorization as Hispanic-other thus provided insufficient information about ancestry and had the potential to confound the study. 4 American Indian, 18 Native Hawaiian and 3 self-identified multiracial without further ancestry delineation were excluded because the numbers of patients in these categories were too few to gather generalizable information. There remained 142 parturitions with maternal African ancestry (self-identified as Black, African or African American), 265 with Asian ancestry (self-identified as Asian) and 1220 with European ancestry (self-identified as White, including Hispanic and non-Hispanic). Of these, RNA sequencing was performed on 9 sPE and 9 normotensive controls of African ancestry, 18 sPE and 15 normotensive controls of Asian ancestry and 23 sPE and 49 normotensive controls of European ancestry (Figure 1).

**Figure 1:**
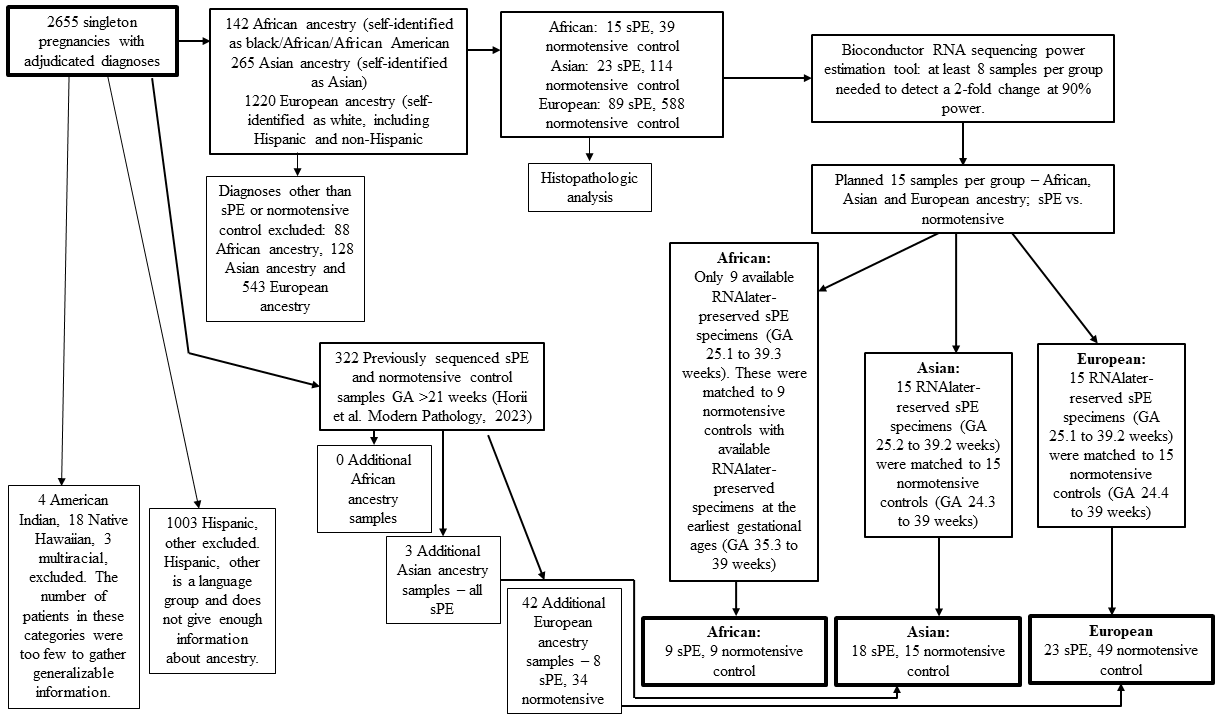
RNA sequencing study design. Abbreviations sPE: preeclampsia with severe features; GA: gestational age

### Obstetric Data

The demographic data collected included maternal ancestry, maternal age at delivery, gravidity, parity, gestational age at delivery, fetal sex, birth weight, mode of delivery, documented sPE, intrauterine growth restriction (IUGR), unexplained tachycardia, or peripartum cardiomyopathy. *RNA isolation from placental tissue, bulk RNA sequencing and analysis* Placenta disc villous tissue (avoiding the chorionic and basal plates) was collected and stored in RNAlater (ThermoFisher). Total RNA was isolated from RNAlater-preserved specimens with the mirVana RNA isolation kit using the manufacturer’s Total RNA protocol (ThermoFisher) and RNA concentration was measured using the QuBit RNA BR assay kit (ThermoFisher). RNA integrity was evaluated using RNA 6000 Nano chip read by a 2100 Bioanalyzer (Agilent). All samples had an RNA integrity number of >7.0. RNA sequencing libraries were prepared using the TruSeq Stranded Total RNA Sample preparation kit with Ribo-Zero Gold (Illumina) at the IGM Genomics Center, UCSD. The libraries were pooled and sequenced on the NovaSeq 6000 (Illumina) with PE100 to an average depth of 25,000,000 uniquely mapped reads. Quality control was performed using FastQC (version 0.11.3). Reads were mapped to GRCh38.89 (Ensembl) using STAR (version 2.7.9a)^17^ on the Extreme Science and Engineering Discovery Environment (XSEDE)^18^. The STAR parameters used were as follows: –runThreadN 24 –sjdbOverhang 99 – genomeDir ref_ensembl_GRCh38_89/star_index –sjdbGTFtagExonParentGenegene_id – sjdbGTFfile ref_ensembl_GRCh38_89/ Homo_sapiens.GRCh38.89.gtf –outSAMtype BAM Unsorted SortedByCoordinate –outReadsUnmapped Fastx –quantMode Gene-Counts. Ensembl genes with at least 10 counts in 25% of the samples were filtered for the analysis. Normalization and the differential expression analysis (DEA) were performed using the R (version 4.2.1) package DESeq2 (version 1.36.0)^19^. Data were accounted for batch effect, gestational age, and gender, and adjusted p-value < .05 was considered differentially expressed. BiomaRt (version 2.42.1) was used to convert Ensembl gene identification numbers to Human Genome Organization gene names^20^. Gene set enrichment analysis (GSEA) was performed using the R (version 4.2.1) package Fast Gene Set Enrichment Analysis (version 1.22.0). Gene ontology analysis was performed using Enrichr^21^. Pathway analysis was performed using MsigDB Hallmark gene set pathway analysis^22^. Venn diagrams were constructed using BioVenn^23^.

### Immunohistochemistry

Formalin-fixed paraffin-embedded placental tissue sections of 5 μm were stained with mouse anti-CD123 (clone BR4MS; 1:100; Ventana) antibody using a manual staining method. Briefly, slides were incubated at 60°C for 1 hour, deparaffinized through standard xylene and alcohol changes and hydrated in distilled water. Slides were pretreated in BioCare BORG decloaking chamber at 125°C, 123psi. Slides were rinsed in TBS (BioCare Medical), peroxidase block (3% hydrogen peroxide) was applied for 10 mins, after which slides were rinsed in TBS. Protein block with 0.5% casein (Vector) was performed for 10 mins, after which slides were rinsed in TBS and then incubated with the CD123 antibody for 30 mins, rinsed in TBS and incubated in post-primary reagent (rabbit anti-mouse Ig; Nvocastra) for 30 mins. Slides were again rinsed in TBS and then Novolink polymer (HRP-polumer-goat-anti-rabbit IG; Leica microsystems) was applied for 30 mins. Slides were rinsed in TBS and then DAB reagent (DAKO) was applied. Slides were rinsed in water, counterstained with hematoxylin, dehydrated, cleared with xylene and then coverslip applied, per standard protocols. Immunohistochemistry was performed at San Diego Pathologist Medical Group.

### Immunohistochemistry quantification

CD123-stained placenta sections were evaluated to by a pathologist; cellular localization was evaluated and scores were assigned based on intensity (no staining = 0, weak staining = 1, moderate staining = 2 and strong staining = 3) and amount of staining (none = 0, focal = 1, patchy = 2 and diffuse = 3). The total score comprised the sum of the intensity and amount scores (lowest = 0 and highest = 6).

### Peripartum cardiomyopathy sub-study

Electronic medical records of the 2655 singleton pregnancies with adjudicated diagnosis (described in the study cohort section above) were searched using the term “cardiomyopathy” and 4 patients with bona fide peripartum cardiomyopathy (PPCM) in the index pregnancy were identified (Figure 6A).

### Statistical analysis

Statistical analyses were performed using GraphPad Prism (version 9.4.1). The Mann-Whitney U test was applied to all nonparametric data and student t test to all parametric data with continuous variables. The Pearson chi-square test was used for categorical variables. All statistical tests were 2-sided, with values <.05 considered statistically significant.

## Results

### Histopathologic analysis cohort

The histopathologic analysis cohort was comprised of 868 patients; 127 with sPE and 741 normotensive controls. Maternal age, gestational age at delivery, gravidity, parity, infant sex, infant birth weight and mode of delivery were evaluated in sPE patients African (n=15), Asian (n=23) and European (n=89) ancestries compared to normotensive controls of African (n=39), Asian (n=114) and European (n=588) ancestries. Gestational age at delivery and birth weight were significantly less in sPE patients than in normotensive controls across ancestries (Supplemental Table 1).

### In patients with severe preeclampsia, African ancestry is associated with higher rates of maternal vascular malperfusion and perivillous fibrin deposition

We analyzed the 868 placentas using previously defined histopathologic criteria^24–25^. We found maternal vascular malperfusion and perivillous fibrin deposition occurred at significantly higher rates in placentas from parturients of African ancestry with sPE compared to those of Asian or European ancestry, suggesting worse placental injury in the African ancestry sPE group (Supplemental Table 2).

### RNA sequencing patient demographics

Maternal age, gestational age at delivery, gravidity, parity, infant sex, infant birth weight and mode of delivery were evaluated in sPE versus normotensive control patients of African, Asian and European ancestries whose placenta samples were RNA sequenced. In comparison to normotensive controls, infant birth weight was significantly lower in sPE parturitions across ancestries while gestational age at delivery, gravidity and parity were significantly less in African ancestry sPE parturitions (Supplemental Table 3).

Among the sPE patients, there was no significant difference in evaluated demographics – maternal age, gestational age at delivery, gravidity, parity, infant sex, birth weight and mode of delivery – across ancestries (Supplemental Table 4).

### Canonical preeclampsia genes and pathways are upregulated in placentas from patients of all ancestries with sPE

We first performed differential expression analysis of sPE vs. normotensive controls within each ancestry, and found 146 genes upregulated in African-sPE, 129 genes in Asian-sPE, and 637 genes in European-sPE placentas. 54 genes were upregulated in common in sPE placentas of all 3 ancestries (Figure 2A; Supplemental Tables 5-7). The relatively small number of differentially expressed genes did not show enrichment of ontology terms using hypergeometric statistical tests. We therefore used our ranked gene list to perform gene set enrichment analysis (GSEA) to look for enriched pathways in our dataset. The leading edge analysis identified the most highly upregulated genes and pathways including those involved in metabolism (*LEP*, *HK2*), hormone regulation (*INHA*, *PAPPA2*), and hypoxia/angiogenesis *(FLT1, FSTL3*, *ENG*, *HTRA4*), which have been previously reported to be upregulated in PE^14–15, 26–32^ (Figure 2B-C; Supplemental Tables 8-10). Based on review of prior publications showing evidence of associations with preeclampsia, 9 of the leading edge genes we identified as upregulated in sPE patients across all ancestries were chosen for further analyses (bolded in Figure 2A, arrows in Figure 2C).

**Figure 2:**
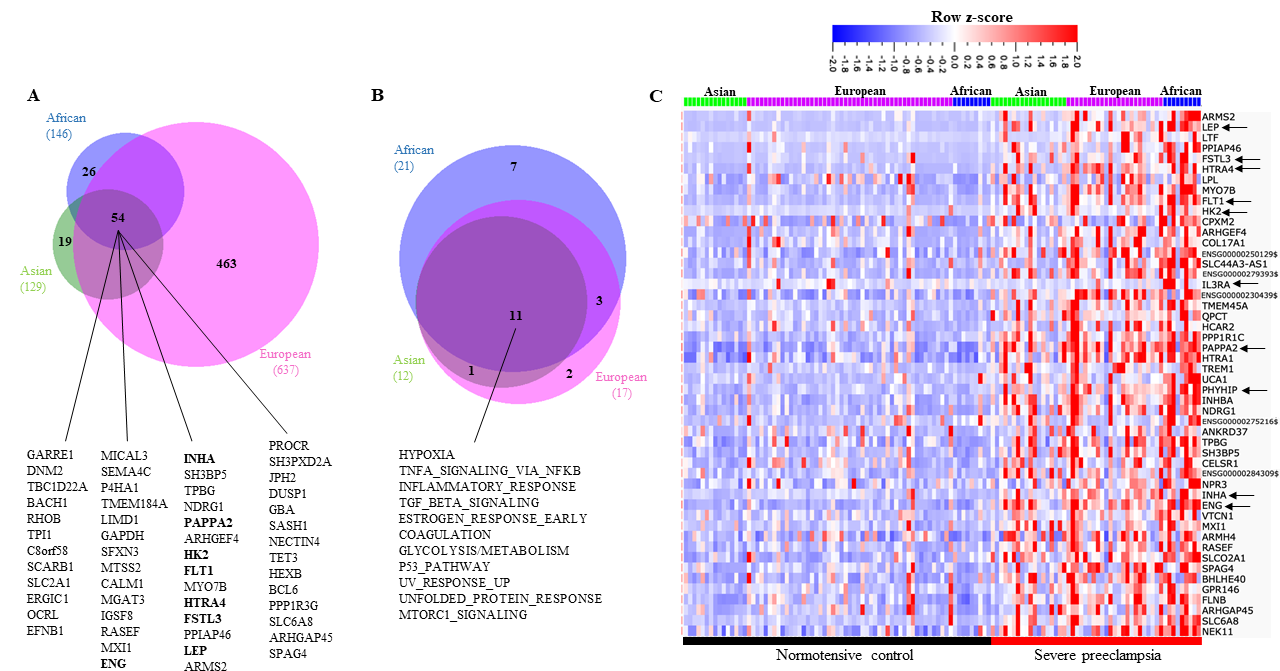
Genes and pathways upregulated in common in placentas from severe PE versus normotensive patients of all ancestries. A. Genes upregulated in severe PE versus normotensive placentas – genes in bold have been identified in multiple studies. B. Hallmark signaling pathways upregulated in severe PE versus normotensive placentas. C. Heat map of top 50 genes upregulated in severe preeclampsia versus normotensive controls. Arrows point to genes chosen for further analysis in this study.

### African ancestry is associated with higher upregulation of PHYHIP and the canonical PE genes, FLT1 and LEP

In evaluating the 9 selected genes upregulated in placentas of sPE patients vs. normotensive controls of all ancestries, we noticed African ancestry sPE placentas showed higher fold-change compared to fold-change from other ancestry groups (Table 1).

**Table 1:** Fold changes in select genes significantly upregulated in severe PE (versus normotensive controls) per ancestry. p values represent adjusted p values calculated using DESeq2.

|  | African ancestry |  | Asian ancestry |  | European ancestry |  |
| --- | --- | --- | --- | --- | --- | --- |
| Gene | Fold | p<sub>adj</sub> | Fold | p<sub>adj</sub> | Fold | p<sub>adj</sub> |
|  | change |  | change |  | change |  |
| ***LEP*** | 35.33 | .0047 | 25.81 | .00058 | 12.70 | 4.61E-06 |
| ***FSTL3*** | 34.03 | .0017 | 11.14 | 3.55E-05 | 2.52 | .0062 |
| ***HK2*** | 9.25 | .0047 | 6.30 | .00058 | 3.97 | 1.26E-05 |
| ***PAPPA2*** | 4.42 | .02 | 4.25 | .0021 | 2.96 | .00015 |
| ***FLT-1*** | 7.54 | .0032 | 5.88 | .0011 | 5.04 | 1.21E-06 |
| ***ENG*** | 4.09 | .0047 | 3.06 | .0095 | 2.66 | 2.48E-05 |
| ***HTRA4*** | 13.56 | .0076 | 6.15 | .027 | 4.96 | 7.49E-06 |
| ***INHA*** | 6.52 | .0367 | 3.38 | .0041 | 2.38 | .00024 |
| ***PHYHIP*** | 6.69 | .0094 | 2.53 | NS | 2.60 | .00038 |

Therefore, we further investigated which of the 9 genes were specifically upregulated in sPE placentas of African ancestry versus sPE placentas of Asian and European ancestries (excluding normotensive controls). We found *FLT1*, *LEP* and *PHYHIP* were more highly upregulated in placentas from African ancestry sPE patients (Figure 3A; Supplemental Figure 1).

**Figure 3:**
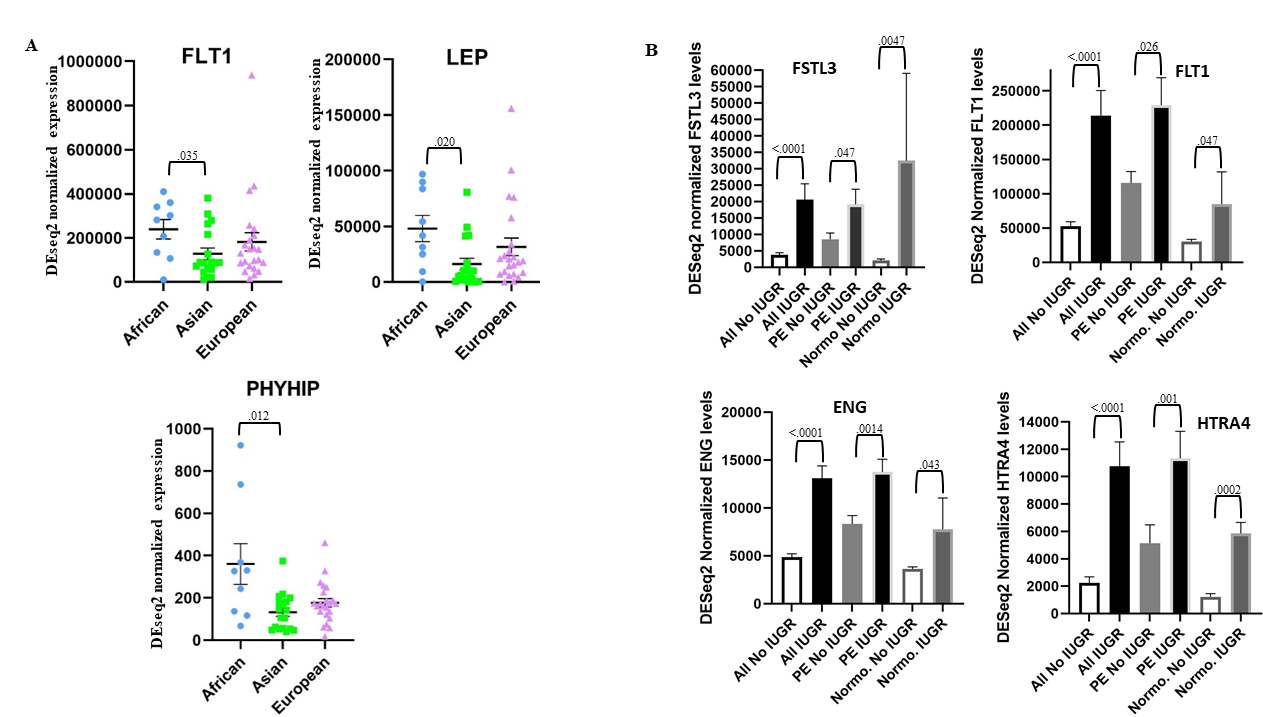
A. DEseq2 normalized expression of FLT1, LEP and PHYHIP which are more highly upregulated in placentas from African ancestry patients with severe preeclampsia versus Asian ancestry patients with severe preeclampsia. B. Evaluation of intrauterine growth restriction (IUGR) in relation to DESeq2 normalized gene expression.

### High placental upregulation of FSTL3 is associated with intrauterine growth restriction

We reviewed electronic medical records of all 123 study patients for intrauterine growth restriction (IUGR), a known PE-associated pregnancy complication. IUGR was present in 28 of 123 patients (6 of 9 sPE African ancestry, 8 of 18 sPE Asian ancestry, 11 of 23 sPE European ancestry, 0 of 9 normotensive African, 0 of 15 normotensive Asian and 3 of 49 normotensive European patients). We evaluated IUGR in relation with DESeq2 normalized gene expression levels of our 9 selected genes. We found that all 9 genes were significantly associated with IUGR when sPE and normotensive IUGR patients were combined. However, only *FSTL3*, *FLT1*, *ENG* and *HTRA4* were associated with IUGR independently of preeclampsia; with *FSTL3* being the most highly upregulated gene (15.4-fold) in normotensive patients with IUGR (Figure 3B; Supplemental Figure 2).

### Allograft rejection and adaptive immune response genes and pathways are selectively upregulated in placentas from patients of African ancestry with sPE

In comparison to placentas from normotensive patients, 26 genes and 7 pathways were uniquely upregulated in placentas from sPE patients of African ancestry and not in Asian or European ancestry sPE placentas (Figure 4A-B). Allograft rejection and adaptive immune response (interleukin-JAK-STAT signaling) pathways were among those uniquely upregulated in placentas from African ancestry sPE patients. Interestingly, leading edge gene analysis identified interleukin receptor, *IL3RA*, which is known for allograft rejection through interleukin signaling^33–35^, to be selectively upregulated in placentas of sPE patients of African ancestry. Therefore, *IL3RA* was chosen for further investigation in our study. We performed regression analysis of *IL3RA* levels in placentas of sPE patients of African, Asian and European ancestries delivered at various gestational ages and found *IL3RA* levels to be elevated in African ancestry sPE placentas, regardless of gestational age at delivery (Supplemental Figure 3A). In African ancestry parturients, individual placentas showed upregulation of both canonical (exemplified by *FLT1* and *LEP*) and allograft rejection/adaptive immune response genes (exemplified by *IL3RA*) (Supplemental Figure 3B).

**Figure 4:**
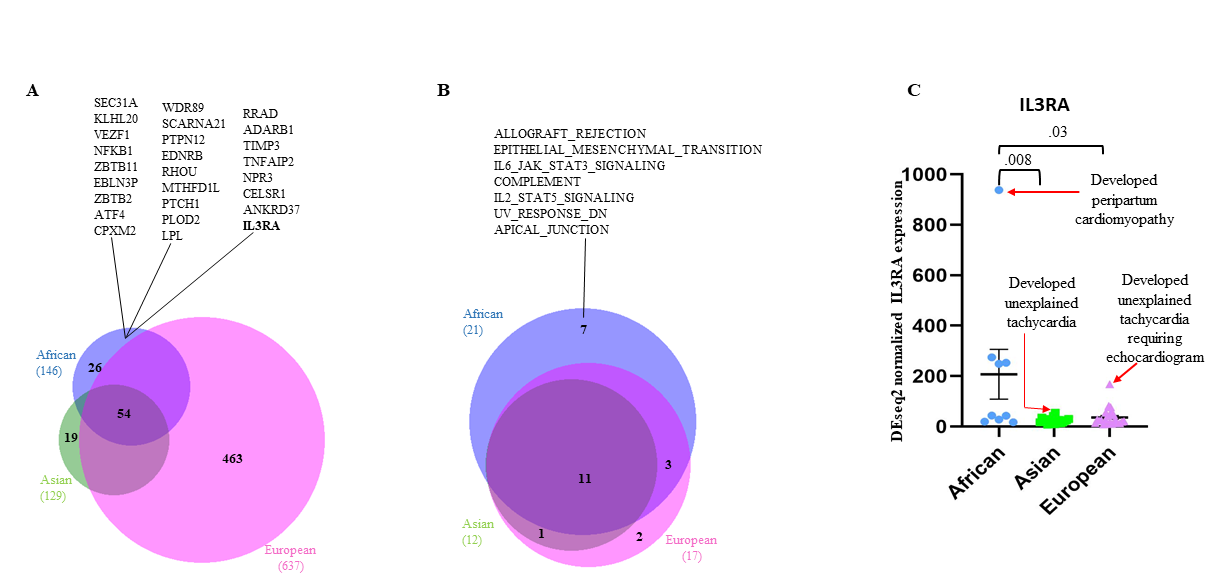
Genes and pathways uniquely upregulated in placentas from severe PE patients of African ancestry and not in Asian or European ancestry severe PE patients. A. Genes uniquely upregulated in placentas from severe PE patients of African ancestry - IL3RA is bolded due to its significance in this study. B. Hallmark signaling pathways uniquely upregulated in placentas from severe PE patients of African ancestry. C. Patients with the highest placental IL3RA levels for their ancestry developed cardiac symptoms.

### High placental upregulation of IL3RA is associated with unexplained tachycardia and peripartum cardiomyopathy

We performed a detailed review of the electronic medical records of the sPE patients with the highest placental *IL3RA* expression levels for their ancestry and found they developed either peripartum cardiomyopathy (PPCM; African ancestry) or unexplained tachycardia (UET; Asian and European ancestries) (Table 2; Figure 4C). Tachycardia in the absence of clinically explained causes such as chorioamnionitis, urinary tract infection, viral infection, post-partum hemorrhage/anemia, dehydration, sleep apnea and bulimia was classified as UET. We thus searched for UET and PPCM in the medical records of all 50 sPE study patients and found elevated *IL3RA* levels to be significantly associated with UET or PPCM (Fisher’s exact test p value .0005) (Table 3).

**Table 2:** Peripartum cardiac symptoms and recurrent preeclampsia in the severe preeclampsia with the highest placental *IL3RA* levels for their ancestry. Abbreviations: PE: preeclampsia, SOB: shortness of breath, IUGR: intrauterine growth restriction, LVEF: left ventricular ejection fraction, LV: left ventricle; EF: ejection fraction

| **Ancestry** | ***IL3RA***<br>**level** | **Fold greater**<br>**than average**<br>**PE levels for**<br>**ancestry** | **Peripartum cardiac symptoms** | **Recurrent PE**<br>**(Y/N)?** |
| --- | --- | --- | --- | --- |
| **African** | 938.05 | 4.5 | Cough, SOB, abdominal distension 5 months post-partum. Found to have dilated cardiomyopathy with 10% LVEF; diagnosed with peripartum cardiomyopathy | Y- severe PE with IUGR 6 years later |
| **Asian** | 54.91 | 2.3 | Unexplained tachycardia that self-resolved; did not require an echocardiogram. | Y – superimposed PE 2 years later |
| **European** | 167.82 | 4.6 | Tachycardia to 120s peripartum – required echocardiogram to exclude peripartum cardiomyopathy; echo showed mild LV diastolic dysfunction and mild pulmonary hypertension, but preserved EF; tachycardia eventually self-resolved | Unknown – had a subsequent pregnancy 1 year later, but no further records after 10w4d visit |

**Table 3:**
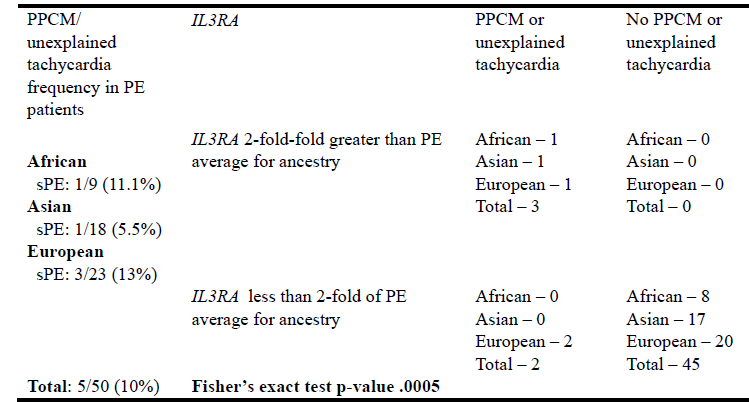
Frequency of unexplained tachycardia or peripartum cardiomyopathy and their relationship with *IL3RA* levels in parturitions complicated by preeclampsia with severe features (sPE) per ancestry (N=50). Comparison placental *IL3RA* levels were set to 2-fold greater than average of the levels in sPE patients per ancestry.

### Placental syncytiotrophoblast CD123 expression by immunohistochemistry correlates with placental IL3RA levels by RNA-seq

We evaluated the localization and expression pattern of CD123, the protein product of *IL3RA* by immunohistochemistry on formalin-fixed paraffin-embedded (FFPE) sections from sPE patient placentas with the highest and lowest placental *IL3RA* levels for their ancestry (i.e. the 2 highest and 2 lowest *IL3RA*-expressing sPE placentas of each ancestry; N =12). There was variable CD123 expression in chorionic plate vessel endothelial cells, stem villous vessel endothelial cells, decidual vessel endothelial cells and syncytiotrophoblasts (Figure 5A-D). CD123 expression in syncytiotrophoblasts correlated with *IL3RA* levels (simple linear regression p = .0027), but expression in stem villous vessels did not (p = .1782) (Figure 5E-F).

**Figure 5:**
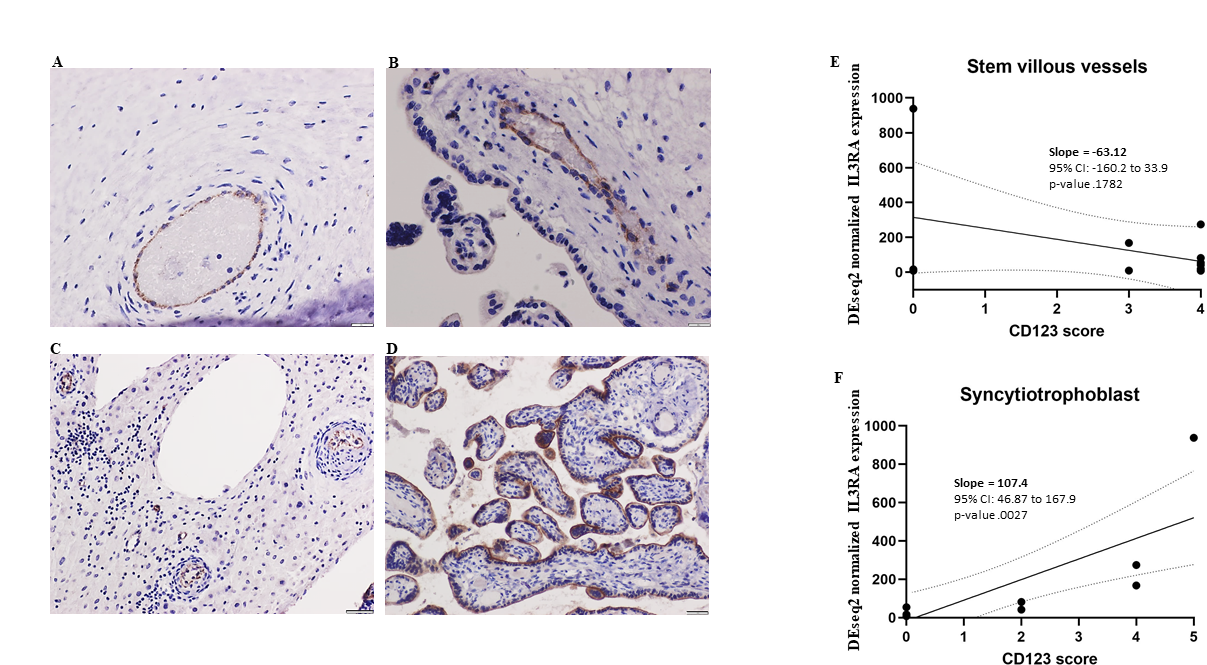
CD123 localization in placenta sections; in placentas with CD123 expression, variable expression was seen in chorionic plate vessels (A), stem villous vessels (B), decidual vessels (C) and syncytiotrophoblasts (D) (top panels are from a patient without PPCM; bottom panels are from the patient with PPCM). CD123 expression in syncytiotrophoblasts correlated better with IL3RA levels than expression in stem villous vessels (E and F). Abbreviations: PPCM peripartum cardiomyopathy; UET: unexplained tachycardia.

### Placental syncytiotrophoblast CD123 expression was detectable in only one of four peripartum cardiomyopathy patients

We reviewed our database of 2655 singleton parturitions to identify patients who developed peripartum cardiomyopathy (Figure 6A; Table 4).

**Figure 6:**
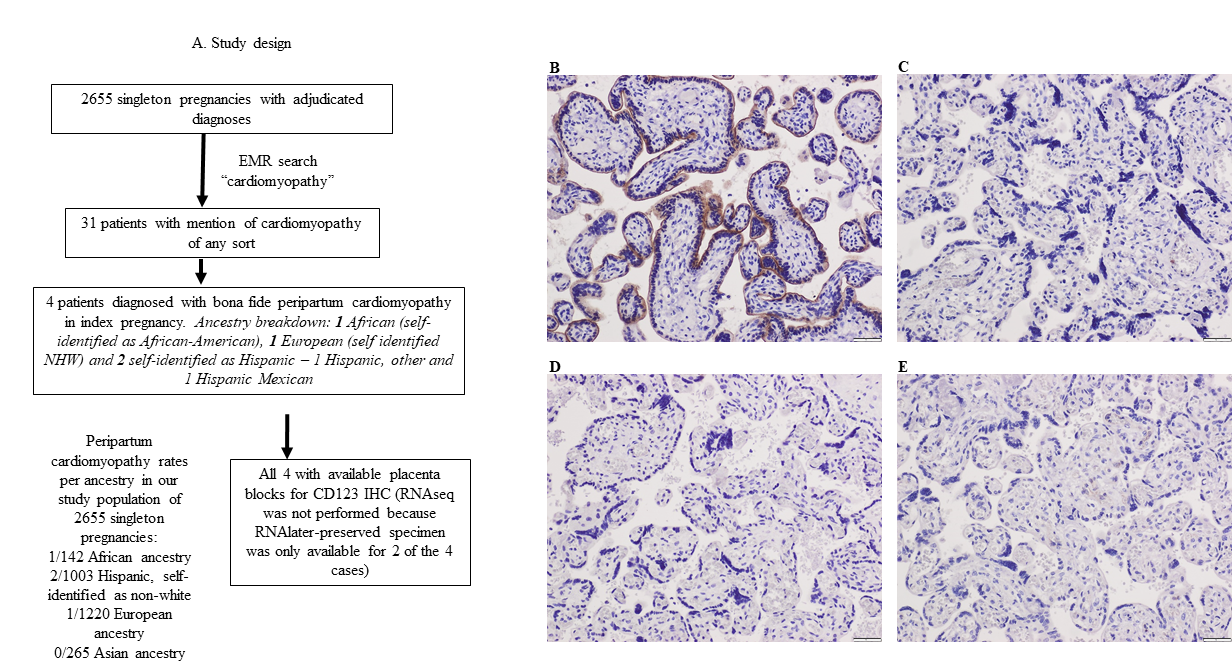
Evaluating the relationship between placental CD123 expression and peripartum cardiomyopathy. A. Study design. CD123 immunohistochemical stains were performed on the PPCM patients of African ancestry (B), European ancestry (C) and Hispanic language group (D-E). Only the African ancestry patient had detectable syncytiotrophoblast CD123 expression.

**Table 4:**
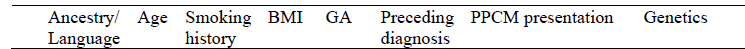
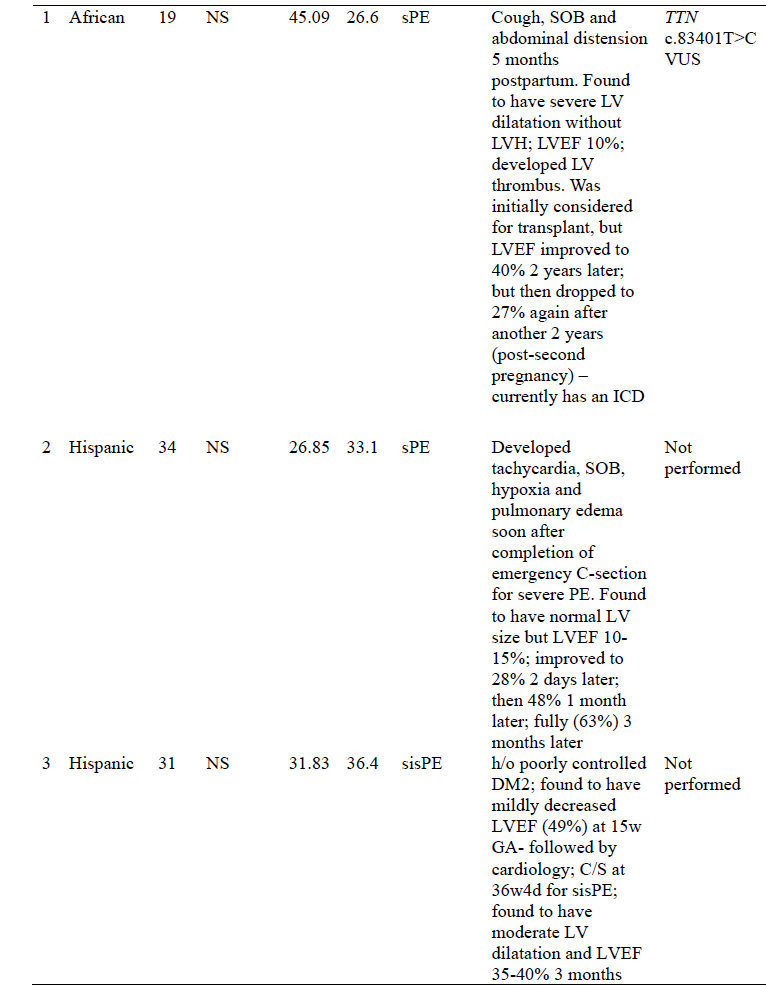
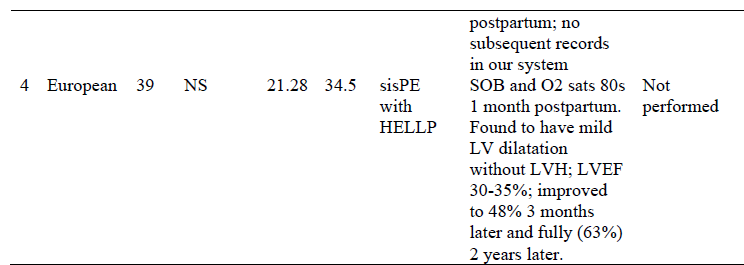
Clinical profile of the 4 patients with peripartum cardiomyopathy (PPCM) in our database of 2655 singleton parturitions. Abbreviations: NS: never-smoker; BMI: body mass index; GA: gestational age; sPE: preeclampsia with severe features; sisPE: superimposed preeclampsia (i.e. preeclampsia with a history of underlying chronic hypertension) with severe features; HELLP: hemolysis, elevated liver enzymes and low platelets; SOB: shortness of breath; LV: left ventricle; LVH: left ventricular hypertrophy; LVEF: left ventricular ejection fraction; ICD: implantable cardioverter defibrillator; DM2: type II diabetes mellitus; VUS: variant of uncertain significance.

We subsequently evaluated placental CD123 expression in the 4 PPCM patients by immunohistochemistry on FFPE tissue sections. Syncytiotrophoblast CD123 expression was present only in the African ancestry PPCM patient (Figure 6B-E).

## Discussion

Factors underlying increased PE risk and worse PE outcomes in African ancestry pregnancies and parturitions are incompletely understood. In the current study, we evaluated placental gene expression by RNA sequencing in patients with sPE of African, Asian and European ancestries and found upregulation of canonical PE genes in sPE patients of all ancestries, with follistatin-like 3 (*FSTL3*) being associated with IUGR. *FSTL3* is an activin antagonist that Mukherjee et al. found to be involved in glucose and fat metabolism^36^. In their unsupervised placental gene expression profiling, Leavey et al^14–15^ found *FSTL3* to be the top gene associated with canonical PE and IUGR. A 2021 study found elevated maternal serum levels of FSTL3 during pregnancy to be predictive of fetal growth restriction^37^. Our findings are in concordance with these studies. Among the genes upregulated in PE across ancestries, we found higher expression of, Fms-related receptor tyrosine kinase 1 (*FLT1*), leptin (*LEP*) and phatonyl-CoA 2-hydroxylase interacting protein (*PHYHIP*) in African ancestry sPE placentas. *FLT1* is probably the most widely recognized PE-associated gene, having been found to be upregulated in PE in multiple studies^26–28, 38^. The ratio of soluble FLT-1 (sFlt-1) to placental growth factor (PIGF) has been validated as a test to predict impending PE^39^. *LEP* plays a major role in the regulation of energy homeostasis and body weight^40^. Elevated *LEP* has been previously described in preeclampsia in multiple studies and leptin infusion in mice induces clinical characteristics of preeclampsia^29–31^. Longitudinal measurement of leptin/ceramide ratio outperformed sFlt-1/PIGF ratio in predicting impending PE^31^. *PHYHIP* has been described to be involved in protein localization and has been suggested to be involved in the development of Refsum disease, a neurologic disorder caused by the toxic accumulation of phytanic acid in brain and peripheral neurons^41^. A few studies have identified *PHYHIP* as one of the placenta genes associated with early-onset preeclampsia^42–43^. Higher upregulation of *FLT1*, *LEP* and *PHYHIP* in placentas from pregnancies and parturitions of maternal African ancestry may partly explain worse PE outcomes in this patient population.

We found genes and pathways involved in allograft rejection and adaptive immune responses to be selectively upregulated in placentas from parturient persons of African ancestry with sPE and not in those of Asian and European ancestries. Notably, individual placentas from patients of African ancestry with sPE had upregulation of both canonical PE genes and allograft rejection/adaptive immune response genes. This raises the possibility that rather than being indicative of distinct PE etiologies (i.e. canonical versus immunologic), activation of allograft rejection/adaptive immune response genes may be a marker of worse placental injury and harbinger of worse PE outcomes.

Among the allograft rejection and adaptive immune response genes, we found high placental expression of interleukin 3 receptor alpha (*IL3RA*) to be associated with unexplained tachycardia and peripartum cardiomyopathy in sPE patients. IL3RA is a cell surface receptor for IL3. Binding of IL3 to IL3RA causes heterodimerization with the beta subunit, IL3RB, and downstream JAK2-STAT5 signaling^44^. IL5 and GM-CSF share the beta subunit with IL3 and can activate signal transduction upon binding^45^. High levels of IL3 and GM-CSF have been observed in various models of heart failure. Vistnes et al.^46^ found IL3 and GM-CSF among the cytokines increased upon inducing heart failure in a mouse cardiomyopathy model. IL3 caused allograft fibrosis and chronic rejection of mouse heart transplants^35^. IL3 impaired the cardioprotective effects of endothelial cell-derived extracellular vesicles in a rat cardiac ischemia/reperfusion model^47^. In humans, Oren et al.^48^ found CRP, GM-CSF and IL3 to be significantly upregulated in the first 12 hours of patients presenting to the emergency department with acute myocardial infarctions.

We found placental endothelial cells and syncytiotrophoblasts express CD123, the protein product of *IL3RA*, with syncytiotrophoblast CD123 expression being a better correlate of *IL3RA* levels by RNA-sequencing. Syncytiotrophoblasts and decidual endothelial cells interact with the maternal systemic circulation at the intervillous space and decidua respectively^49–50^. CD123/IL3RA localization to these cell types suggests *IL3RA* may exert its adverse cardiovascular effects via interactions with the maternal systemic circulation and circulating immune cells. In support of this theory is the finding by Anzai et al.^51^ that T-cell derived IL3 acted on *IL3RA*-expressing cardiac macrophages and fibroblasts to amplify autoimmune inflammation in a mouse experimental myocarditis model. Furthermore, Pistulli et al^52^ found accumulation of CD123+ dendritic cells in hearts of humans with acute myocarditis, and that dendritic cell accumulation caused adverse left ventricular remodeling in murine model of experimental myocarditis. IL5, which shares the signal transducing beta subunit with IL3-IL3RA, is a contributor to the pathogenesis of eosinophilic myocarditis^53–56^. All together, the upregulation of placental *IL3RA* in patients with sPE who develop unexplained tachycardia and/or peripartum cardiomyopathy suggest that this marker may be a predictor and/or cause of cardiac dysfunction, perhaps via interaction with circulating immune cells that then accumulate in, and cause damage to, the heart. Studies of the crosstalk between syncytiotrophoblasts, immune cells, and cardiomyocytes may yield further mechanistic insights into how the placenta may cause cardiovascular damage in susceptible patients.

The patient with the highest placental *IL3RA* expression level in our RNA sequenced cohort of 123 patients developed peripartum cardiomyopathy (PPCM). PPCM is a non-ischemic dilated reduced ejection fraction (LVEF <45%) cardiomyopathy that typically presents towards the end of pregnancy and in the months following delivery^57^. The incidence of PPCM varies by ancestry, being highest among people of African ancestry, with incidence rates reaching 1:100 in Nigeria^58^, and lowest among people of Asian ancestry, with rates as low as 1:20,000 in Japan^59^. Currently, the etiology of PPCM is thought to be multifactorial, with proposed causes that include genetic predisposition in 15% of PPCM patients^60^, vasculotoxicity of a 16kDa cleaved prolactin fragment^61^, and inflammation^62^. There is an increased association of PPCM with hypertensive disorders of pregnancy, with the association being strongest with sPE^63^. A 2018 study showed a higher prevalence of loss of function variants in cardiomyopathy genes among patients with PE; with *TTN* mutations being most common^64^. Interestingly, the African ancestry sPE patient in our study who had the highest placental *IL3RA* level and developed PPCM also had a c.83401T>C variant of uncertain significance (VUS) in *TTN*.

We subsequently performed a review of our database of 2655 singleton parturitions and identified 3 additional PPCM patients (4 total, including the African ancestry patient whose placenta had been RNA sequenced). Of the 4 patients, syncytiotrophoblast CD123 expression, our surrogate marker for high placental *IL3RA*, was identified in 1 and absent in 3. A possible explanation for this finding includes differing PPCM etiologies in the 4 patients. The 3 patients without evidence of elevated *IL3RA* were older at delivery (age range 31-39 years), and recovered cardiac function with normalization of their LVEF by 1-2 years postpartum. In contrast, the patient with elevated *IL3RA* and *TTN* c.83401T>C was age 19 at delivery and still has decreased LVEF and an implanted ICD 7 years postpartum. Though assessment for cardiomyopathy-associated mutations were not performed in the 3 PPCM patients whose placentas were CD123-negative (due to lack of available preserved RNA), their recovery of cardiac function/normalization of LVEF suggests decreased likelihood of underlying cardiomyopathy-associated genetic mutations. In the IPAC (Investigations of Pregnancy-associated Cardiomyopathy) cohort, the presence of TTN truncating variants significantly correlated with lower LVEF at 1 year follow up^60^.

Using a genomics-first approach to evaluate for heart disease associated with titin-truncating variants, dilated cardiomyopathy was identified in only 7.5% of people with TTN truncating variants^65^. As >90% of patients with TTN truncating variants do not develop dilated cardiomyopathy or PPCM, our findings of elevated placental *IL3RA* in an African ancestry PPCM patient with *TTN* c.83401T>C raises the possibility that high placental *IL3RA* precipitates PPCM in genetically susceptible patients.

Our study was retrospective and limited by the small number of PPCM patients. Prospective studies of placental *IL3RA* expression relative to cardiomyopathy-associated mutation status in large PPCM cohorts are thus warranted.

## Conclusion

We sought to identify placental factors that may explain worse PE outcomes in pregnancies and parturitions with maternal African ancestry and found higher upregulation of canonical PE genes (*FLT1*, *LEP* and *PHYHIP*) and selective upregulation of allograft rejection/adaptive immune response genes (*IL3RA*) in placentas from sPE patients of African ancestry versus sPE patients of Asian and European ancestries. Furthermore, we found high placental *IL3RA* to be associated with UET and PPCM in a subset of sPE patients. The higher expression of *LEP*, *FLT1*, *PHYHIP* and *IL3RA* in placentas from patients of African ancestry with sPE likely contributes to the worse PE outcomes in this patient population. As pregnancy is a semi-allogeneic state and HLA diversity is higher in people of African ancestry^66^, the possibility that the higher upregulation of canonical PE genes, and selective upregulation of allograft rejection-associated genes in pregnancies and parturitions with maternal African ancestry may be related to higher rates of maternal-fetal HLA mismatches in this population, is an intriguing concept for future studies.

## Data Availability

RNA-seq data have been deposited to the Gene Expression Omnibus database under the accession number GSE234729 and GSE186257.

## Acknowledgement

The authors are very grateful to all patients who consented and donated placental tissues to our Perinatal Biorepository. We also thank Dr. Mana Parast for constructive critiques on the research and manuscript.

## Author contributions

OA and MH designed the research; CM and MM recruited patients and collected clinical information; MBJ and MM collected and managed database; LLS, and LCL were involved in review of data within the obstetrics registry and adjudication of hypertensive disorders of pregnancy; OA and HSL adjudicated adverse outcomes; TB performed RNA isolation; OA and DP performed immunohistochemistry analysis; OA, KMF, RM and MH performed RNAseq analysis; OA, MBJ, KMF, LCL and MH wrote the manuscript text. All the authors reviewed the final version of the manuscript.

